# Random effects modelling vs logistic regression for the inclusion of cluster level covariates in propensity scores for medical device and surgical epidemiology

**DOI:** 10.1101/2022.02.02.22269820

**Authors:** Mike Du, Albert Prats-Uribe, Sara Khalid, Daniel Prieto-Alhambra, Victoria Y Strauss, Sara Khalid

**Affiliations:** Nuffield Department of Orthopaedics, Rheumatology and Musculoskeletal Sciences, Botnar Research Centre, Nuffield Orthopaedic Centre, University of Oxford, Oxford, UK

**Keywords:** propensity score, simulation, trial emulation, clustered data, random effects model, causal inference

## Abstract

**Purpose:** Surgeon and hospital related features such as surgeries volume can be associated with treatment choices and treatment outcomes. Accounting for these covariates with propensity score (PS) analysis can be challenging due to clustered nature of the data. Previous studies have not focused solely on the PS estimation strategy when treatment effects are estimated using random effects model(REM). We studied PS estimation for clustered data using REM compared with logistic regression.

**Methods:** Six different PS estimation strategies were tested using simulations with variable cluster-level confounding intensity (odds ratio(OR)=1.01 to OR=2.5): i) logistic regression PS excluding cluster- level confounders; ii) logistic regression PS including cluster-level confounders; iii) same as ii) but including cross-level interactions; iv), v) and vi), similar to i), ii) and iii) respectively but using REM instead of logistic regression PS. Same analysis were tested in a randomised controlled trial emulation of partial vs total knee replacement surgery. Simulation metrics included bias and mean square error (MSE). For trial emulation, we compared observational vs trial-based treatment effect estimates.

**Results:** In most simulated scenarios, logistic regression including cluster-level confounders gave more accurate estimates with the lowest bias and MSE. E.g. with 50 clusters x 200 individuals and confounding intensity OR=1.5, the relative bias= 10% and MSE= 0.003 for (i), compared to 21% and, 0.010 for (iv). In the Trial emulation, all 6 PS strategies gave similar treatment effect estimates.

**Conclusions:** Logistic regression including patient and surgeon/hospital-level confounders appears to be the preferred strategy for PS estimation. Further investigation with more complex clustered structure is suggested.

**Competing interests:** Prof. Prieto-Alhambra’s research group has received grant support from Amgen, Chesi-Taylor, Novartis, and UCB Biopharma. His department has received advisory or consultancy fees from Amgen, Astellas, AstraZeneca, Johnson, and Johnson, and UCB Biopharma and fees for speaker services from Amgen and UCB Biopharma. Janssen, on behalf of IMI-funded EHDEN and EMIF consortiums, and Synapse Management Partners have supported training programs organised by DPA’s department and open for external participants organized by his department outside submitted work.

**Ethics Approval and Informed Consent:** This study was approved by the secretary of state, having considered the recommendation from the Confidentiality Advisory Group (CAG reference: 17/CAG/0174). Informed ethical approval was given on the use of pseudonymised patients data included in the study.

## INTRODUCTION

Observational studies using routinely collected patient data from health registries are often used for clinical treatment comparative study when randomised control trials are unfeasible or unethical(1). Conversely to randomisation in trials, treatment allocation in observational data is often driven by patient and physician features, leading to confounding by indication. First proposed by Rosenbaum and Rubin(2, 3), propensity score (PS) weighting are a popular method to minimise the resulting bias. Most PS applications in pharmacoepidemiology include only patient covariates. Conversely, medical device and surgical studies typically have a clustered structure that accommodates hospital and physician/surgeon features that could impact treatment and outcome and hence act as confounders(4, 5).

Several simulation studies have shown that using random effects models(6) in the PS estimation or treatment outcome modelling can reduce the bias arising from cluster level confounding in clustered data(7-12). However, it is unclear whether random effects models should be used for both PS estimation and outcome modelling in observational studies of medical devices or surgical procedures. Therefore, this study aims to evaluate to what extent random effects model should be used for PS estimation when random effects model is used to estimate the treatment outcome. This study aimed to assess to what extent random effects model should be used for clustered observation studies of medical device and surgical epidemiology.

We used Monte Carlo simulations(13), and a surgical trial emulation study comparing partial and total knee replacement surgery to evaluate the accuracy and precision of random effects model compared to logistic regression propensity score model.

## METHODS

### Simulation data generation process

The simulation settings were based on previous simulation studies(7, 8) but with parameters adapted to medical device/surgical epidemiology data. We simulated clustered datasets via Monte Carlo simulations with a fixed sample size of 10,000 individuals to represent the patients, binary treatment allocation (T) and binary outcome (Y). The complete mathematical formulae for data generation are included in the supplementary material. We simulated six individuals-level covariates (x1 to x6), two cluster-level covariates (z1 and z2 to represent the hospital/surgeon level covariates) and a cross-level interaction term between the individual and cluster-level confounder for each individual. Among the individual covariates simulated, 5 were confounders (x1- x5), 1 was a risk factor associated with outcome but not with exposure (x7), and 1 was an instrumental variable (x6). Both cluster level covariates (z1 and z2) were generated as confounders. Figure 1 gives the clustered causal diagram of the simulation covariates.

**Figure 1:**
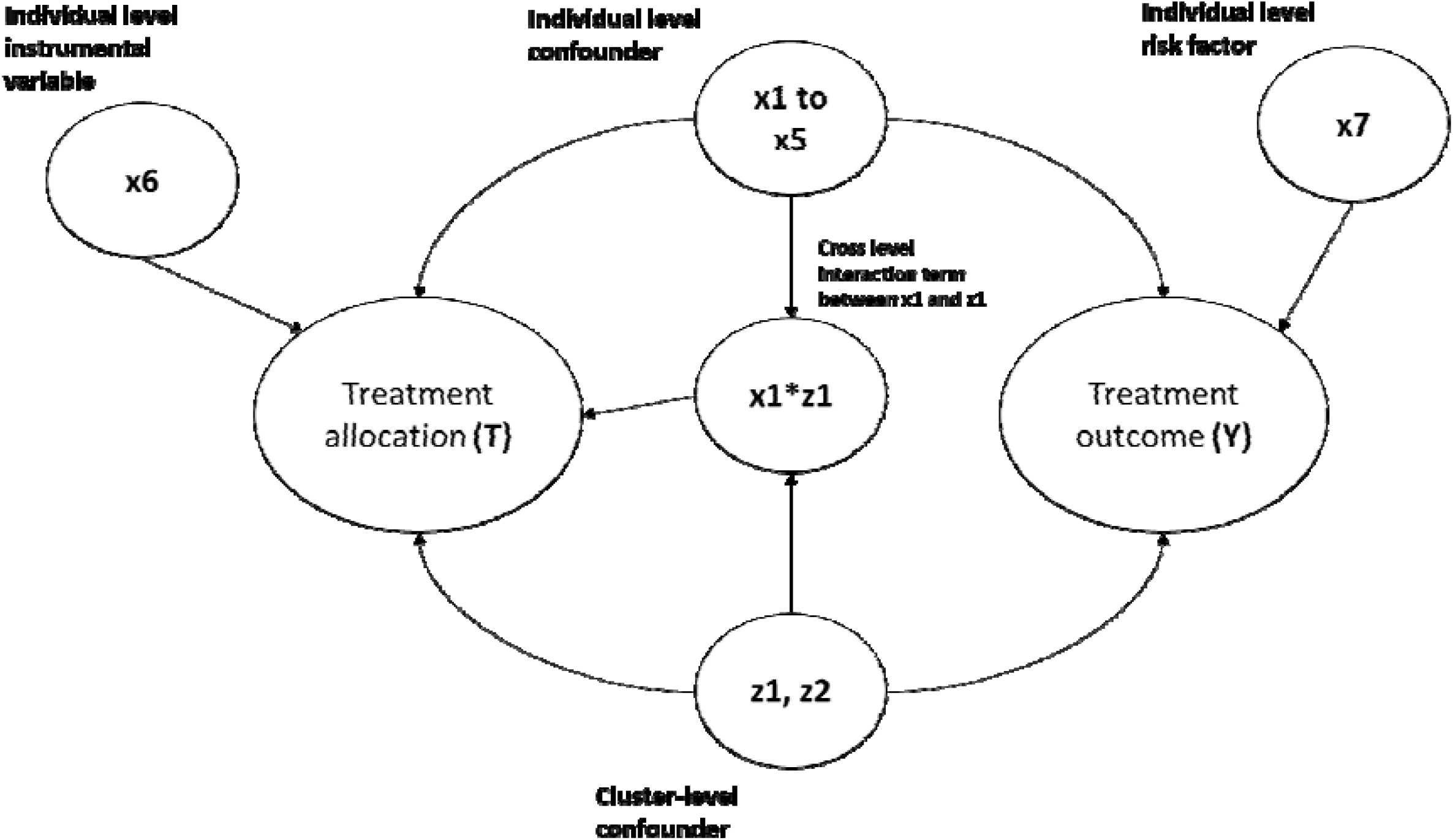
This diagram gives the causal relationship between the covariates in the simulation data, the arrow indicates causes. For example, x1-> Y implies x1 causes Y.

**Figure 2:**
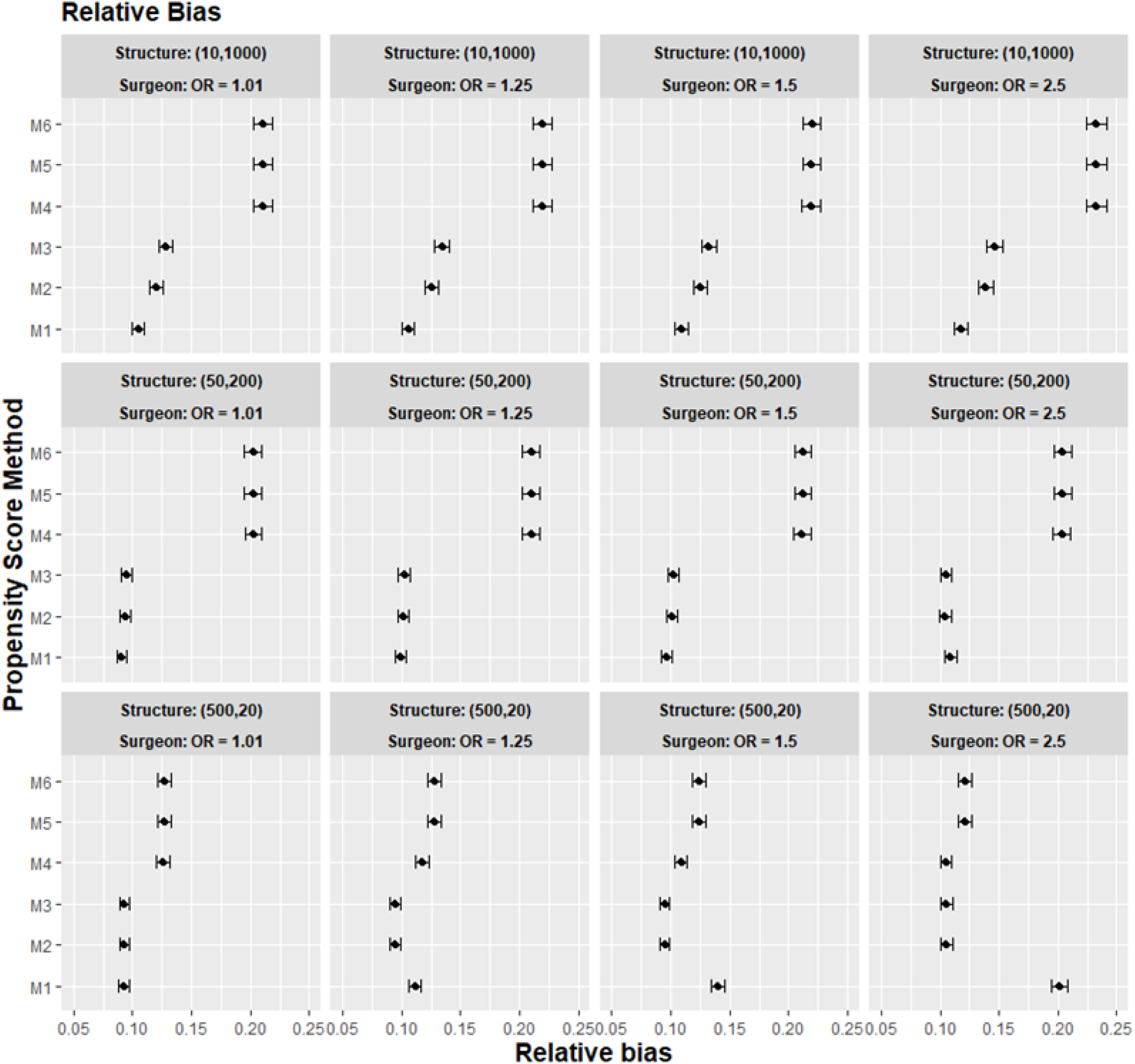
The graphs give the simulation treatment effects average absolute relative bias and 95% confidence interval for propensity score specification strategies M1 to M6 for different cluster structure and cluster (surgeon) level confounder odd ratio on treatment outcome. Structure = (number of clusters, individuals per cluster), surgeon OR = cluster level confounder odd ratio on treatment outcome. Propensity score(PS) strategies: M1 = logistic regression PS excluding cluster-level confounders; M2 = logistic regression PS including cluster-level confounders, M3 = logistic regression PS with cluster level confounders and cross level interaction term, M4 = random effects PS excluding cluster-level confounders, M5 = random effects PS including cluster-level confounders, M6 = random effects PS with cluster level confounders and cross level interaction term

12 different scenarios with 1000 replications under each scenario were simulated to test: 1) three different ratios of cluster: individual size: 10: 1000, 50: 200, and 500: 20; 2) different effect size for z1 and z2 on outcome, ranging from negligible with odds ratio = 1.01 (resembling an instrumental variable) to strong with odds ratio = 2.5 (equivalent to strong multilevel confounding). Table 1 gives the generation distribution, effects on treatment allocation and effects on treatment outcome for all the covariates generated in the simulations.

**Table 1:** The table gives the generation distribution, effects on treatment allocation and effects on treatment outcome for covariates generated in the simulations. OR = odd ratio

| Covariates | Description | Effects on treatment allocation (beta value) | Effects on treatment outcome (Beta value) | Generation distribution |
| --- | --- | --- | --- | --- |
| **z1,z2** | Cluster-level confounders | $z1 = z2 = 0.4055$<br>(equivalent to OR=1.5) | $z1 = z2 =$<br>[0.01,0.2231,0.4055,0.9163]<br>~ [equivalent to OR<br>=1.01,1.25,1.5,2.5] | $z1 \sim N(0,1),$<br>$z2 \sim \text{Bernoulli}(0.5)$ |
| **x1 to x5** | Individual level confounders | $[x1,x2,x3,x4,x5] =$<br>[0.35,0.4,0.45,0.5,0.55] | $[x1,x2,x3,x4,x5] =$<br>[0.35,0.4,0.45,0.5,0.55] | $[x1,x2,x3] \sim \text{Bernoulli}$<br>([0.4,0.45,0.5] )<br>$x4,x5 \sim N(0,1)$ |
| **x6** | Individual level risk factor | 0 | 0.5 | $\text{Bernoulli}(0.5)$ |
| **x7** | Individual level instrumental variable | 0.5 | 0 | $\text{Bernoulli}(0.5)$ |
| **z1\*x1** | Cross level interaction term | 0.4055 (OR =1.5) | 0 | $z1*x1$ |

The simulation data were generated with simstudy (version 0.2.1) R package, and the propensity score models were fitted with lme4 (version 1.1.21) R package.

### Propensity score estimation strategy

For all the data scenario described in the simulation data generation process, we tested six different strategies to incorporate propensity score, as defined in Table 2.

**Table 2:** The table gives the cluster level information contained and the statistical models used for the six propensity score estimation strategies (M1 to M6).

| **Propensity score strategy** | **Cluster level confounders as covariates in PS model** | **Cross level confounders interaction term as covariate in PS model** | **Statistical model to build a propensity score** |
| --- | --- | --- | --- |
| **M1** | Excluded | Excluded | Logistic regression |
| **M2** | Included | Excluded | Logistic regression |
| **M3** | Included | Included | Logistic regression |
| **M4** | Excluded | Excluded | Random effects model<sup>1</sup> |
| **M5** | Included | Excluded | Random effects model<sup>1</sup> |
| **M6** | Included | Included | Random effects model<sup>1</sup> |

<sup>1</sup> The random effects model were built with logit link function

### Treatment effect estimation

For each of the scenarios, the average treatment effect (ATE) was estimated using random effects models with logit function regress on treatment outcome weighted with stabilised inverse probability weighting (SIPW)(14) based on propensity scores calculated using the strategies described in table 2. Random effects model was use for treatment effect estimation since several simulation studies on propensity score(7, 8) have shown that using random effects models to account for the cluster level confounding generally gives the least bias.

### Assessment of simulation results

We measured each propensity score specification strategy’s performance on each scenario by calculating the 1) absolute relative bias (%), defined as the average percentage difference between the true treatment effect and the estimated treatment effect. 2) mean square error (MSE), which is a measure of accuracy. 3) 95% confidence interval model coverage, defined as the proportion of the 95% confidence intervals of the estimated treatment effect effects containing the true treatment effect. All the performance measures were calculated following the simulation study guidelines discussed in Morris et al(15) with “rsimsum” (version 0.9.1) R package.

### Case study on medical device and surgical epidemiology

We used data from the UTMOST study(16), which aimed to identify the optimal methods for controlling confounding when emulating the results of the TOPKAT surgical trial(17). The UTMOST cohort study included patients with a first primary total knee replacement (TKR) or unicompartmental knee replacement (UKR)(18) in the UK National Joint Registry (NJR) from 2009 to 2016 who would have met the TOPKAT trial eligibility criteria. UTMOST included a total of 294556 patients (294556 UKR and 21,026 TKR patients), and 6420 different lead surgeons carried out the interventions. UTMOST extracted 18 patient-level covariates from the NJR, linked to Hospital Episode Statistics (HES) records and patient-reported outcome measures, and the volume of UKR performed by each lead surgeon in the previous year from the NJR. The UTMOST study outcome was revision five years after surgery. Table 3 gives the covariates adjusted in the study.

**Table 3:**
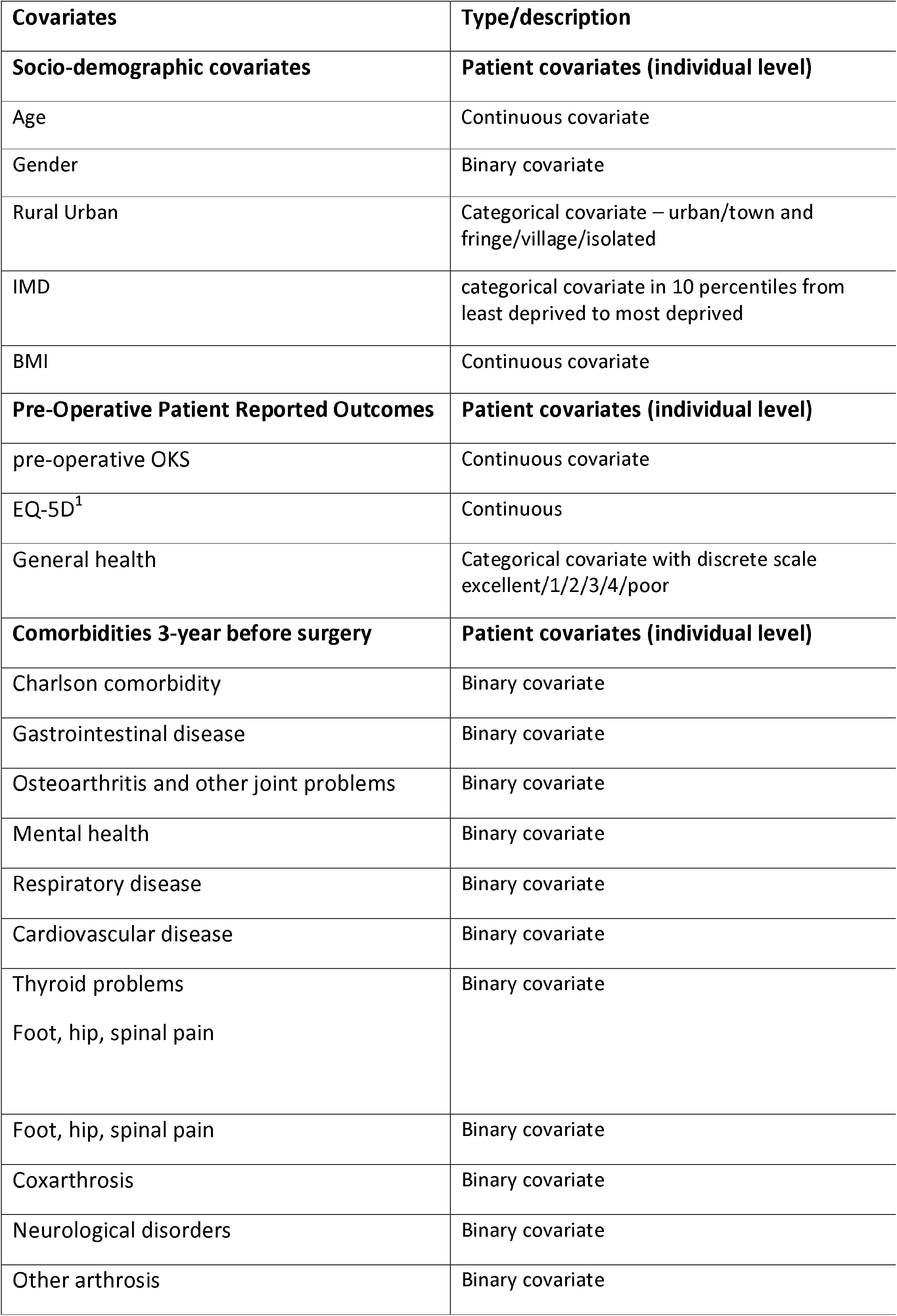
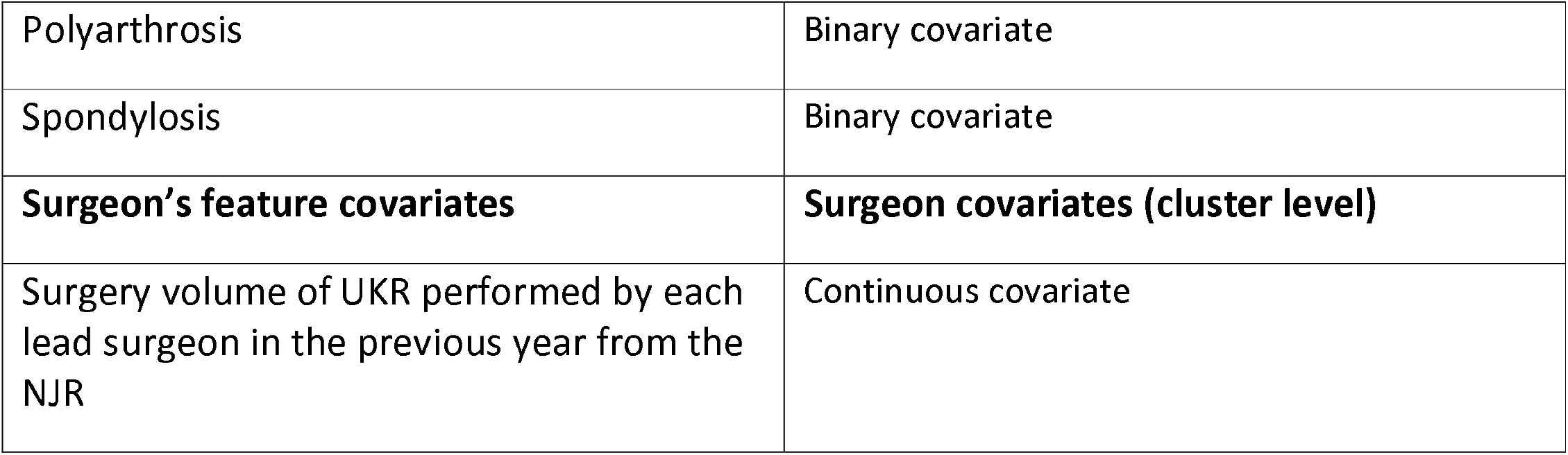
This table gives the covariates adjusted in the case study. UKR = unicompartmental knee replacement NJR = National Joint Registry. 1 standardised measure of health-related quality of life developed by the EuroQol Group

We applied the six proposed propensity score specification strategies from table 2 to the UTMOST dataset to construct the propensity scores for UKR and compared it to the results of the TOPKAT surgical trial. The cross-level interaction term considered in UTMOST was the interaction of surgeon volume and patient gender. As with the simulated data described in the method section, we modelled the 5-year revision risk for patients received UKR using a random effects model with the lead surgeon as cluster level while covariates were adjusted with stabilised inverse probability weights.

## RESULTS

### Simulation study

Figures 3 and 4 gave the simulations average absolute relative bias and MSE of the treatment effect estimates for propensity score estimation strategy M1 to M6. There were few clear trends appeared from figure 3 and 4 that were consistent in all cluster structure scenario. The relative bias and MSE for models with and without the cross-level interaction were similar, for example, relative bias = 9.42% in M2 and relative bias = 9.53% in M3 for cluster level confounders odd ratio (OR) = 1.01, cluster structure (10,1000) scenario, suggesting that not incorporating the cross-level correlation where there is one did not impact on bias much. In scenarios where the cluster level confounders had minimal effect on outcome (OR = 1.01), the model where propensity score without cluster level confounders in the logistic regression (M1) gave the lowest bias when compared to other PS models. By contrast, M1 did not always gave lowest relative bias and MSE when the effect size of cluster level confounders OR were greater than 1.01.

**Figure 3:**
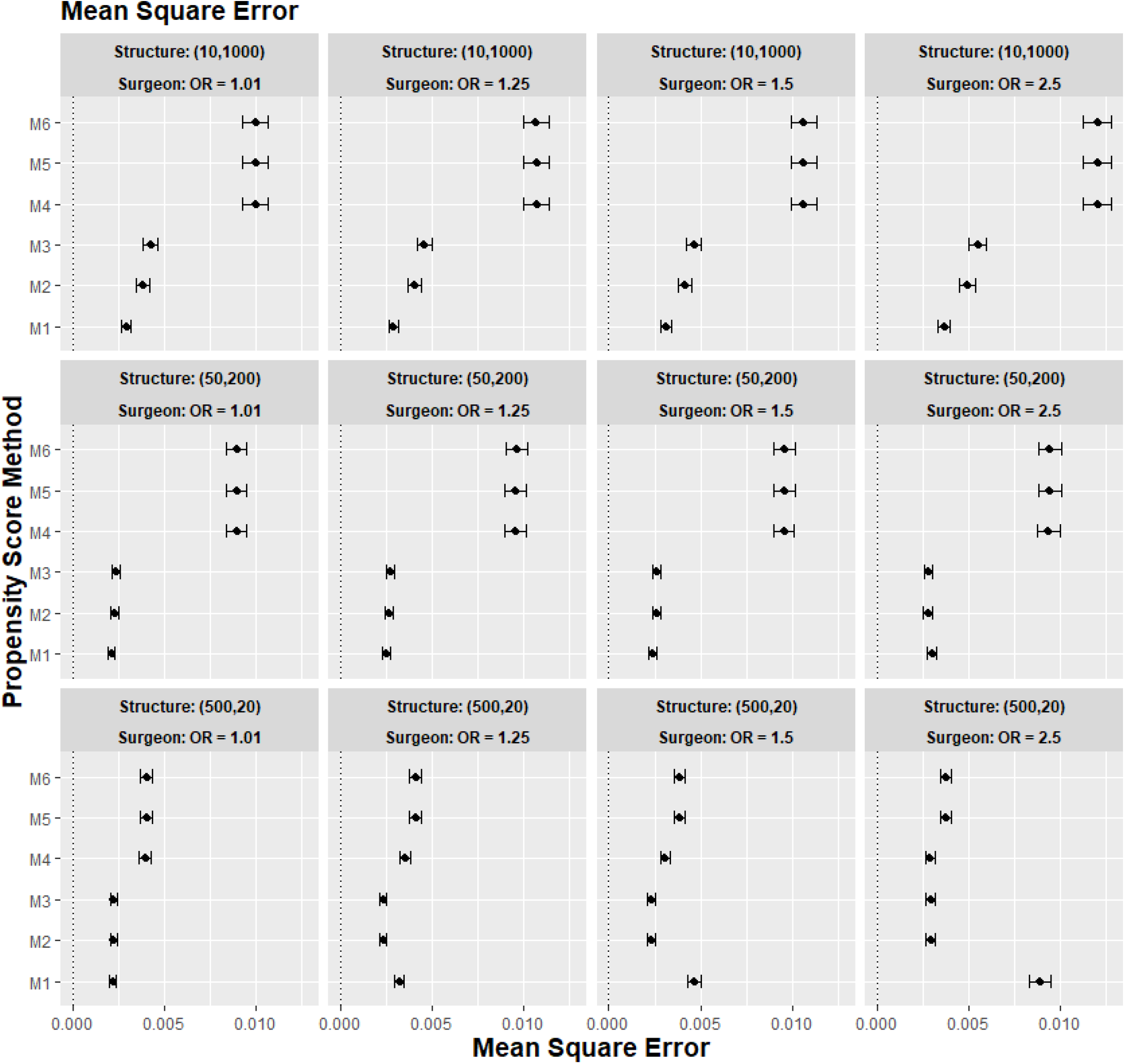
The graphs give the simulation treatment effects average mean square error and 95% confidence interval for propensity score specification strategies M1 to M6 for different cluster structure and cluster (surgeon) level confounder odd ratio on treatment outcome. Structure = (number of clusters, individuals per cluster), surgeon OR = cluster level confounder odd ratio on treatment outcome. Propensity score(PS) strategies: M1 = logistic regression PS excluding cluster-level confounders; M2 = logistic regression PS including cluster-level confounders, M3 = logistic regression PS with cluster level confounders and cross level interaction term, M4 = random effects PS excluding cluster-level confounders, M5 = random effects PS including cluster-level confounders, M6 = random effects PS with cluster level confounders and cross level interaction term

**Figure 4:**
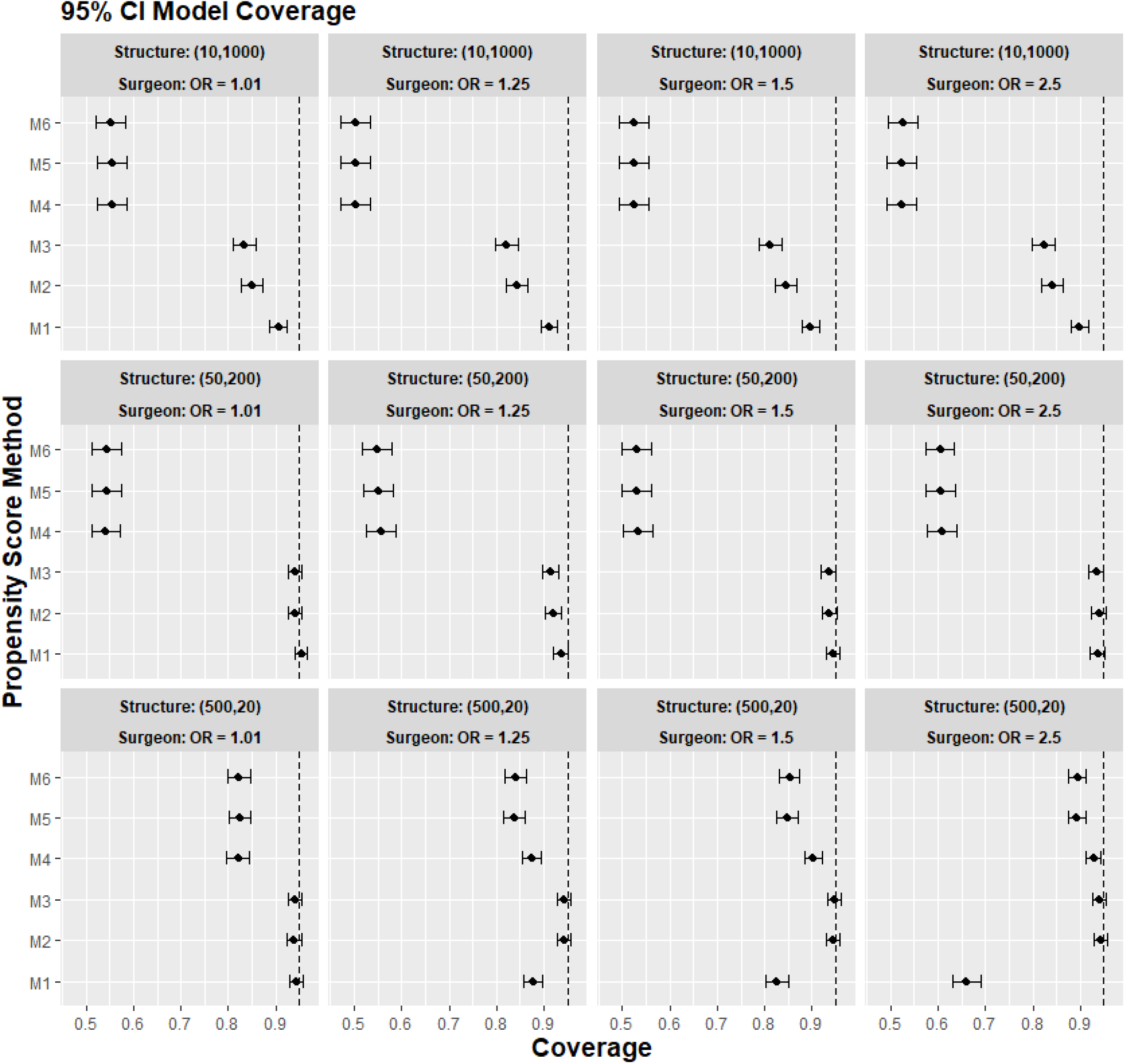
The graphs give the simulation treatment effects average 95%CI model coverage probability and its 95% confidence interval for propensity score specification strategies M1 to M6 for different cluster structure and cluster level confounder odd ratio on treatment outcome. Structure = (number of clusters, individuals per cluster), surgeon OR = cluster level confounder odd ratio on treatment outcome. Propensity score(PS) strategies: M1 = logistic regression PS excluding cluster-level confounders; M2 = logistic regression PS including cluster-level confounders, M3 = logistic regression PS with cluster level confounders and cross level interaction term, M4 = random effects PS excluding cluster-level confounders, M5 = random effects PS including cluster-level confounders, M6 = random effects PS with cluster level confounders and cross level interaction term. The black vertical dotted line indicates 95%.

For cluster structure with small cluster number and large cluster size (m = 10, n = 1000) and (m =50, n =200) using random effects model for propensity score estimation (M4, M5, M6) consistently gave higher bias compared to using logistic regression model (M1, M2, M3). For example, the relative bias for M4 is 21.1% compared to 9.53% for M1 in cluster structure (m = 50, n = 200) and cluster level confounders effect size odd ratio 1.5 scenario. Also, adding the cluster level confounders as covariates in the propensity score model did not impact the bias much in cluster number (m), cluster size (n) = [(10,1000), (50,200)] scenarios regardless of the cluster level confounder effect size on the treatment outcome. Since the relative bias for M1 compared to M2 and M3, and the relative bias M4 compared to M5 and M6 are very similar.

The results for the smallest cluster size scenarios (m = 200, n = 50) behaved differently compared to the other two cluster structures tested in the study. Apart from in the cluster confounder effect on outcome OR = 1.01 scenario. The relative bias for propensity score strategy that either included the cluster level confounders as covariates in the propensity score model or used a random effects model to account for the cluster structure of the data (M2 to M6) reduced bias compared to propensity score strategy did not consider the cluster level (M1). The improvement in bias and MSE was greater as the cluster level confounders effect on outcome increases. For example, the relative bias for M1 = 14.02% compared to M2 = 9.48% for cluster level confounders effect on outcome OR = 1.5. For cluster level confounders effect size on outcome OR = 2.5, the relative bias for M1 = 20.16% compared to M2 = 10.54%.

Figure 4 gave the 95% CI model coverage for the simulation study. It showed that low coverage was a major issue for treatment effects estimates using random effect model based propensity score model (M4 to M6) when the cluster size was large (n = 1000 and n = 200) as the model coverage for M4 M5 M6 were much lower than M1 M2 M3. In our small cluster size scenario (n = 20), the model coverage between M4 M5 M6 and M1 M2 M3 were more closely matched. However M2 and M3 still gave higher model coverage then M4 M5 M6.

### Real world case study

Figure 5 gives the treatment effect estimates using the six propensity score strategies (M1 to M6) proposed for the case study (UTMOST) and the TOPKAT surgical trial estimates. We found that under all model strategies, UKR had a higher risk for 5-year revision than TKR. In contrast, TOPKAT found no statistically significant difference in the revision risk between UKR and TKR. Models that incorporated multilevel data or not or/and included the cluster-level confounders in the propensity score model had an overlapping confidence interval of outcome estimates. This meant all six proposed propensity score strategies (M1 to M6) gave similar treatment estimates and were not statistically significantly different. In addition, propensity score models with and without cross level interaction term had similar estimates (M2 vs M3, M5 vs M6), suggesting that adding the cross level interaction term in the propensity score models did not impact the estimate.

**Figure 5:**
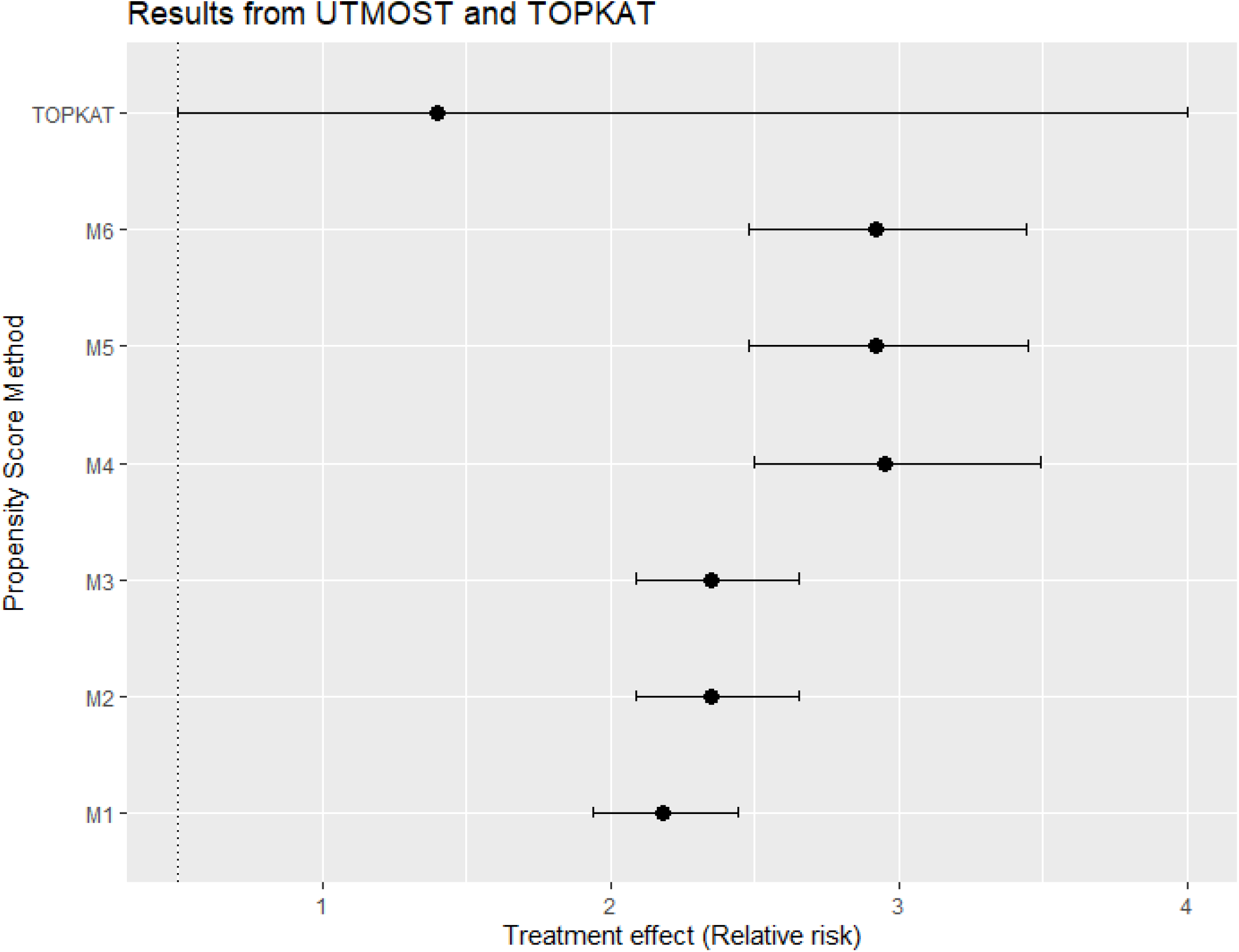
Treatment effects estimates in relative risk and its 95% confidence interval using data from the UTMOST study and the six proposed propensity score strategies M1 To M6 and also the TOPKAT surgical trial estimates. TOPKAT = surgical trial estimates. Propensity score(PS) strategies: M1 = logistic regression PS excluding cluster-level confounders; M2 = logistic regression PS including cluster-level confounders, M3 = logistic regression PS with cluster level confounders and cross level interaction term, M4 = random effects PS excluding cluster-level confounders, M5 = random effects PS including cluster- level confounders, M6 = random effects PS with cluster level confounders and cross level interaction term

## CONCLUSION AND DISCUSSION

### Discussion

This study aimed to find the best way to account for cluster level confounding in propensity score model for propensity score weighting analysis when random effects model was used to estimate the treatment outcome. In our simulation study, we found accounting for the cluster level confounders in the propensity score model when random effects model was used as the outcome model does not always give the smallest bias. For cluster structures with small cluster number and large cluster size (m = 10, n = 1000) and (m =50, n =200), strategy that ignored the cluster level confounders (M1) performed the best. Including the cluster level confounders in the propensity score model by using random effects model and as covariates in the model only offered improvement in bias for small cluster size scenarios (m = 500, n = 20). This is consistent with previous studies on propensity score for clustered data(7, 8, 12), which shows random effects model might give more accurate estimation in propensity score compared to logistics regression but not necessary improvement in accuracy for treatment estimation. However in our simulation study we also showed the optimal propensity score model strategy were dependent on clustered structure and cluster level confounder effect on outcome. Whereas previous simulations study(8, 12) on this topic were more focus on the performance of different weighting approaches. We also found that adding the cross-level interaction term made little impact to the treatment effect in the simulation study.

Applying the proposed propensity score strategies to real-world clinical study corroborated with some but not all our simulation results. Including a cross-level interaction term in either the logistic regression or random effects model did not substantially change the estimated treatment effect, same as the simulation study result. However, the treatment effect estimates in the real-world clinical study all had overlapping confidence intervals, meaning all six propensity score strategies (M1 to M6) gave similar results, different from our simulation results. There were few differences between the cluster structure, which could contribute to these differences in the result. First, the cluster size was fixed in the simulation study, but the cluster size was varied for the real-world clinical study. Second, we found that many surgeons only carried out one type of treatment in the real-world clinical study. However, in our simulation study, the treatment is allocated individually, meaning both treatments can appear in all clusters. This discrepancy of results between our real-world clinical study and simulation also highlighted that the cluster structure of the data affects the accuracy and precision of results for propensity score weighting analysis. More research is needed on how different cluster structures affect propensity score weighting analysis.

### Strengths and Limitation of the study

This study’s main strength is its use of both simulations and real-world data. Using simulated data, where the true average treatment effect was known, allowed us to compare the accuracy of the six proposed PS estimation strategies. Using clinical data allowed us to test whether the trends from the simulation study were held with real-world data.

This study has several limitations. In the simulation study, we investigated only fixed cluster number and size scenarios in the scenarios. Our real-world case study found the simulation findings may not be able to generalise to the scenario when cluster number and size was not fixed. In addition, we only tested the propensity score strategies on binary outcomes. Therefore, our results cannot generalise to other types of outcomes. We also assumed that the treatment assignment was only influenced by a small set of covariates in the simulation study. It could be argued that in real world settings, the data would usually contain more covariates. However, the focus of this study was not on covariates number. Finally, the TOPKAT trial treatment estimate was underpowered in the real-world case study. As a result, the 95% confidence interval for the trial treatment estimate was large, making it difficult to compare the accuracy of the treatment effects from the UTMOST data.

### Conclusion

In summary, careful consideration of the cluster structure is necessary to decide on whether to use a random effects model on propensity score estimation. We should only consider using random effects model for propensity score model when the dataset contains large numbers of small clusters. Also, we should consider including cluster level confounders as covariates in the propensity score model when the cluster level confounders are thought to strongly affect the treatment outcome, as this can reduce bias.

## Data Availability

All data produced in the present study are available upon reasonable request to the correspondence author Prof Daniel Prieto-Alhambra

## Abbreviations

PS: propensity score;
OR: odd ratio;
REM: random effects model;
MSE: mean square error;
UKR: unicompartmental knee replacement;
TKR: total knee replacement;
NJR: UK National Joint Registry

