## supplemental for "Random effects modelling vs logistic regression for the inclusion of cluster level covariates in propensity scores for medical device and surgical epidemiology"

**Supplementary material**

A. **A detailed description of the simulation generation process for the simulation study**

Data generation with coefficients are defined below

Let consider a two-level data structure for our scenario in which the patient level (1st level) is indexed as i where i =1, 2, …….. , Nj. Then I are nested under surgeon level (2nd level) indexed by j (where j = 1, 2, 3, ….J)

Let T the treatment allocation be defined as:

$$T_{\left\{ \mathrm{ij} \right\}}\sim Bernoulli\left( \pi_{\left\{ \mathrm{ij} \right\}} \right)$$

$$\log\frac{\left\{ 1-\pi_{\left\{ ij \right\}} \right\}}{\left\{ \pi_{\left\{ ij \right\}} \right\}} = b0+c0_{j}+ b1x1_{\left\{ ij \right\}}+ b2x2_{\left\{ ij \right\}}+ b3x3_{\left\{ ij \right\}}+ b4x4_{\left\{ ij \right\}}+ b5x5_{\left\{ ij \right\}}+ b6x6_{\left\{ ij \right\}}+ b7z1_{\left\{ j \right\}}+ b8z2_{\left\{ j \right\}}+ b9x2_{\left\{ ij \right\}}*z1_{\left\{ j \right\}}$$

where $c0_{j\sim}N\left( 0,0.5 \right)$

Let Y the clinical outcome be defined as:

$$Y_{\left\{ ij \right\}}\sim Bernoulli\left( \alpha_{\left\{ ij \right\}} \right)$$

$$\log\frac{\left\{ 1-\alpha_{\left\{ \mathrm{ij} \right\}} \right\}}{\left\{ \alpha_{\left\{ \mathrm{ij} \right\}} \right\}}= d0+e0_{j}+ d1x1_{\left\{ \mathrm{ij} \right\}}+ d2x2_{\left\{ \mathrm{ij} \right\}}+ d3x3_{\left\{ \mathrm{ij} \right\}}+ d4x4_{\left\{ \mathrm{ij} \right\}}+ d5x5_{\left\{ \mathrm{ij} \right\}}+ d6x7_{\left\{ \mathrm{ij} \right\}}+ d7z1_{\left\{ j \right\}}+ d8z1_{\left\{ j \right\}}+ B* \mathrm{Tr}t_{\left\{ \mathrm{ij} \right\}}$$

where $d0_{j\sim}N\left( 0,0.5 \right)$and Trt is the treatment allocation. All other variables are definite in the table below. All the variables are independent.

| Variable name | Variable Type | Distribution/Value Assigned |
| --- | --- | --- |
| z1,z2 | Surgeon level confounder | z1 = normal(0,1), z2 = Bernoulli(0.5) |
| z1*x1 | Cross level interaction term | z1*x1 |
| x1, x2, x3, x4, x5 | Patient-level confounder | x1,x2,x3=Bernoulli ([0.4,0.45,0.5] )  x4,x5 = normal(0,1) |
| x6 | Patient-level instrumental variable | Bernoulli( 0.5) |
| x7 | Patient-level risk factor | Bernoulli( 0.5) |
| b1 to b5 | Beta values for patient level confounder on treatment allocation | = [0.35, 0.4, 0.45, 0.5, 0.55] |
| b6 | Beta values for patient level instrumental variable | = 0.5 |
| b7,b8 | Beta values for surgeon level confounder on treatment allocation | b7 = 0.4055, b8 = 0.4055 |
| b9 | Beta value for cross level interaction term | = 0.4055 |
| b0 | fixed intercept on treatment allocation | = -0.5 |
| c0j | c1 is the random surgeon effect on treatment allocation (intercept) | Normal(0, 0.5) |
| d1 to d5 | Beta values for patient level confounder on outcome | = [0.35, 0.4, 0.45, 0.5, 0.55] |
| d6 | Beta values for patient level risk factor | = 0.5 |
| d7 | Beta values for surgeon level confounder on outcome | = [0.01, 0.2231, 0.4055,0.9163]= odd ratio (1.01,1.25,1.5,2.5)  to test performance different strategy for IPW |
| d0 | fixed intercept on outcome | = -0.5 |
| e0j | the random surgeon effect on outcome (intercept) | Normal(0, 0.5) |
| B | True treatment effect | 0.4055 equivalent to odd ratio 1.5 |
| N | Total sample size | Fixed at 10000 |
| (m,n) | Cluster number, cluster size | = (10,1000),(50, 200), (500,20) |
